# Universal properties of the dynamics of the Covid-19 pandemic

**DOI:** 10.1101/2020.08.24.20181214

**Authors:** Piotr T. Chruściel, Sebastian J. Szybka

**Affiliations:** Faculty of Physics, University of Vienna; Astronomical Observatory, Jagiellonian University; Copernicus Center for Interdisciplinary Studies

## Abstract

We present evidence for existence of a universal lower bound for the initial growth rate of the epidemic curve of the SARS-CoV-2 coronavirus. This can be used to infer that, on average, an asymptomatic infected individual is infectious during 5.6 ± 0.3 days. We further present evidence of an average time scale of 12 days for halving the number of new cases, or new deaths, during the extinction period of the first phase of the epidemic.

## 1 Introduction

The development of strategies to fight future phases of the Covid-19 pandemic, or similar future epidemics, requires a good understanding of the global features of the evolution of the epidemic. In particular it is important to obtain quantitative information about the epidemic which is independent of the local conditions such as social habits of the local population or the details of the confinement measures. In this work we show existence of a universal lower bound on the rate of the initial growth of the epidemic, with a characteristic time scale of about 6 days. We argue that this minimal rate of growth can be explained by an infectiousness time of infected individuals of 6 days. Furthermore, we show that the epidemiological data exhibit an average extinction rate with a characteristic time scale of 12 days. The information should be useful for monitoring the development of the current epidemic for countries which are still in the first phase thereof, as well as that of an upcoming second wave, or future coronavirus epidemics.

Our results are based on fitting to the epidemiological time-series the curves

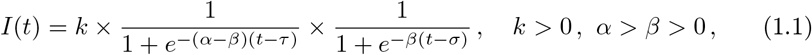

introduced in [1] (see also Appendix C), which describe surprisingly well the global features of the confirmed-cases time-series and death time-series for the first phase of the Covid-19 epidemic, whenever a fit to the data is available. Here one should keep in mind that in many countries the epidemic stabilised to a new rampant phase, or entered a new growing phase, in which case the fits of the function *I* to the data are becoming worse with time. This allows us to determine, for each time-series, the end of the first phase of the epidemic defined as the day for which the fit of the curve is optimal.

A shortcoming of the curve (1.1) is that *I* can only provide a good fit for data sets for which the extinction rate is smaller than half of the initial growing rate; this is explained in Appendix A. While this is a fundamental bias of our analysis, it turns out that many Covid-19 time-series have this property.^1^

A key feature of the function *I* is that if both *e*^(^*^α^*^−^*^β^*^)^*^τ^* and *e^βσ^* are sufficiently large,^2^then near *t* =0 the function *I* is well approximated by

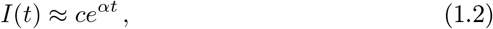

where *c* = *ke*^−(^*^α^*^−^*^β^*^)^*^τ^*^−^*^βσ^*. This shows that the parameter *α* carries information about the *initial rate of growth* of the epidemic.

Next, for large positive *t* the function *I* behaves as

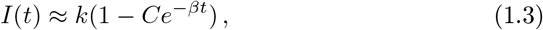

when ever *C* = *e^βσ^* or *t* (or both) are sufficiently large. This shows that *β* describes the *extinction rate* of the epidemic.

Fitting the curve to the time series of the epidemic for a given city, county, province, state, or country, provides quantitative information about the dynamics of the epidemic.

We have attempted to determine the parameters (*α, β, k, τ, σ*) for all time-series of the epidemic available at the John Hopkins University (JHU) server on July 1, 2020. As already pointed-out in [1], *optically satisfactory* fits to the time series of the curve (1.1) can be found for essentially all time-series that we have examined; several examples can be found below, in[1], and in the supplementary materials to this paper. However, it appears that *satisfactory* fits, in the sense of *small residuals and decent standard deviations for the fit parameters*, can typically only be found after the (first) peak of the number of new cases of epidemic. Moreover, wobbly time-series, or time series with jumps, often do not lead to fits which are satisfactory in this sense either. While our observations below are based on the time series with satisfactory fits, we checked that they are consistent with all fits when the large uncertainties of the parameters of the remaining fits are taken into account.

The values of all fitting parameters depend upon the country considered for many obvious reasons, such as awareness of the epidemic, quarantine measures taken, social habits of the local population, etc. The main observation of this work is the existence of absolute lower bounds for the constants *α* and *β*, independently of local factors. As reported in more detail in Section 2, we find the following lower bounds for all time-series examined:

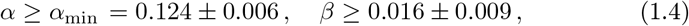

with the minimum for *α* attained at the time series of the county Cumberland, New Jersey in the US, and the minimum for *β* attained for the State of Victoria in Australia. This is illustrated in Figure 1_1, with the fits for Cumberland and Victoria shown in Figures 1.2 and 1.3. (Unless explicitly indicated otherwise, error indicators are standard deviations.)

**Figure 1.1:**
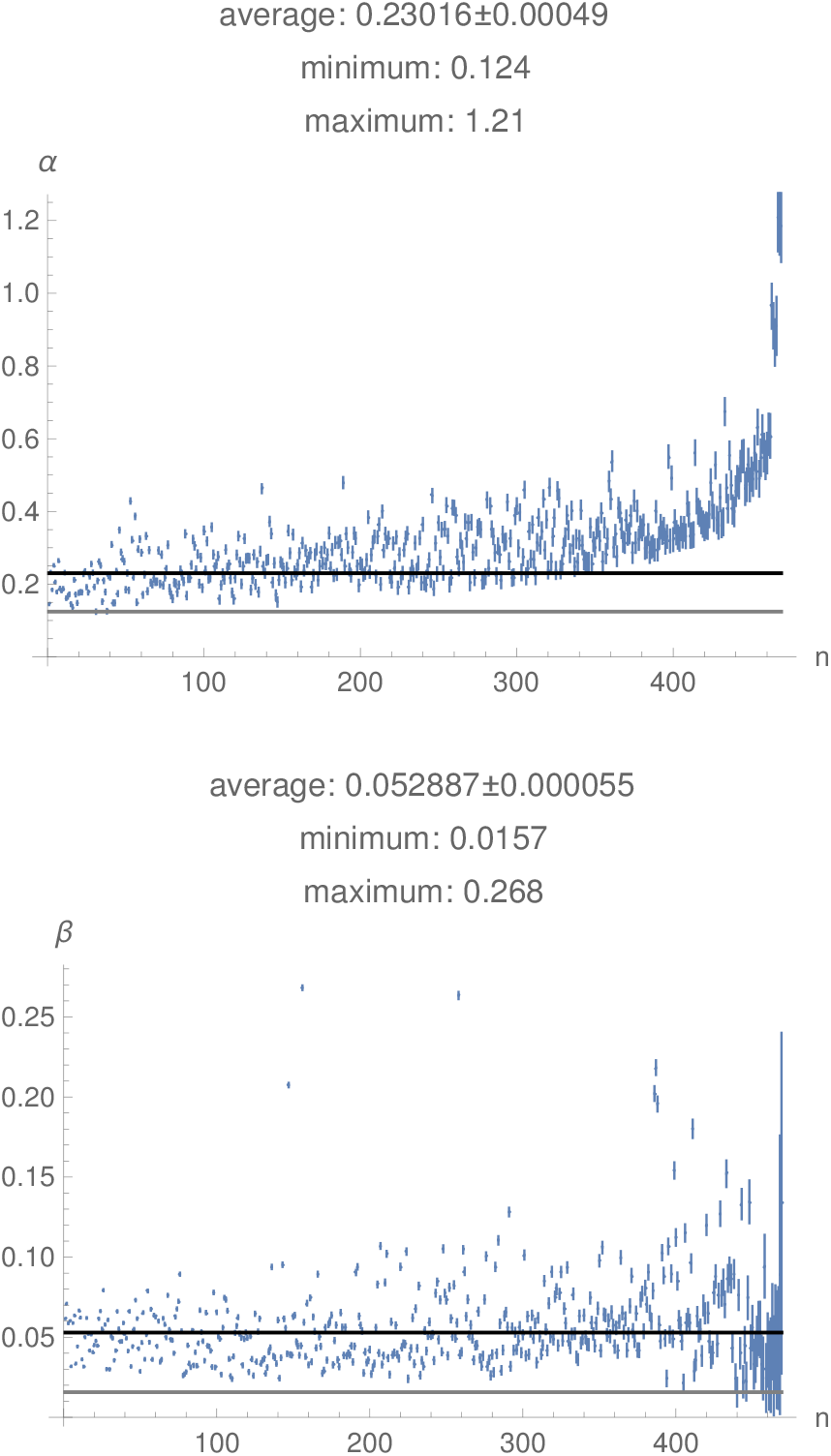
The values of the parameters *α* and *β* for the confirmed-cases time series, ordered by increasing standard deviation. Only those fits where all relative errors of the fit are less than 10% were used in the analysis and are shown. The black line is the mean and the grey line is the minimum value.

**Figure 1.2:**
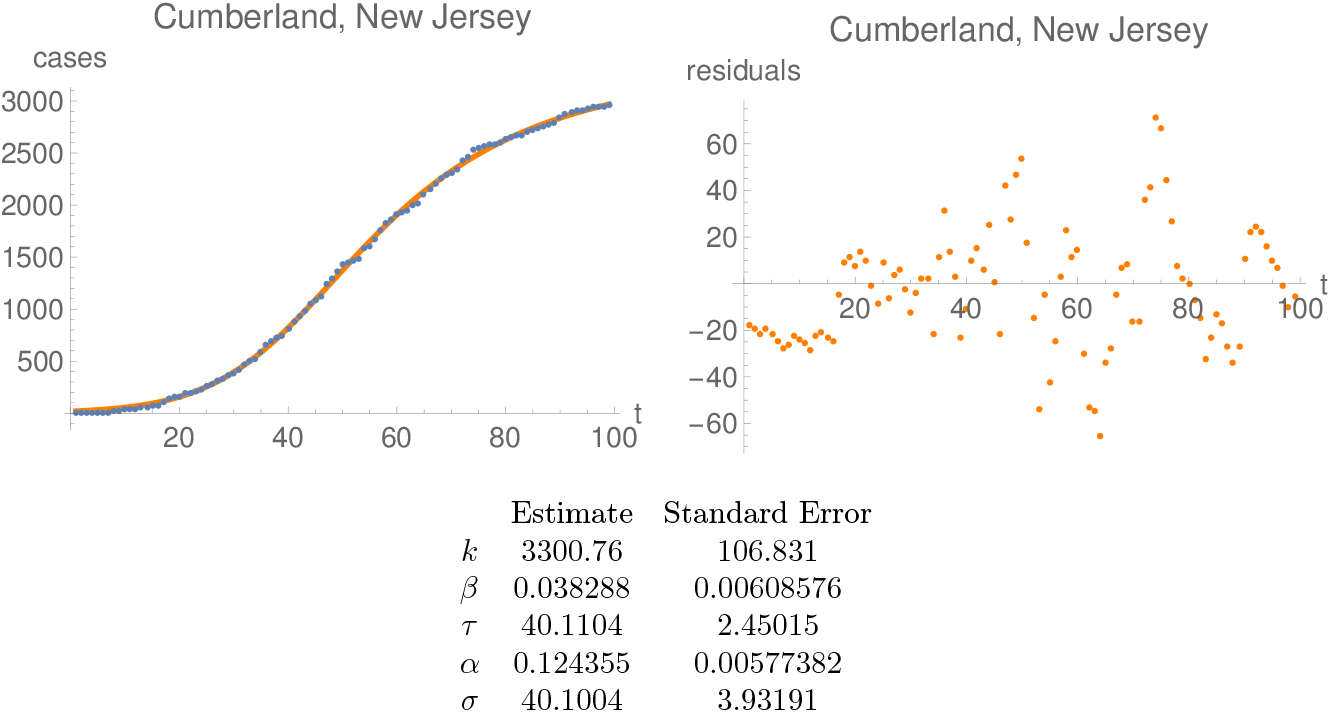
Plots of the time series and the best fit (left) before June 30, 2020, as well as the residuals of the fit (right) for the confirmed-cases time-series with smallest *α*, namely Cumberland, New Jersey.

**Figure 1.3:**
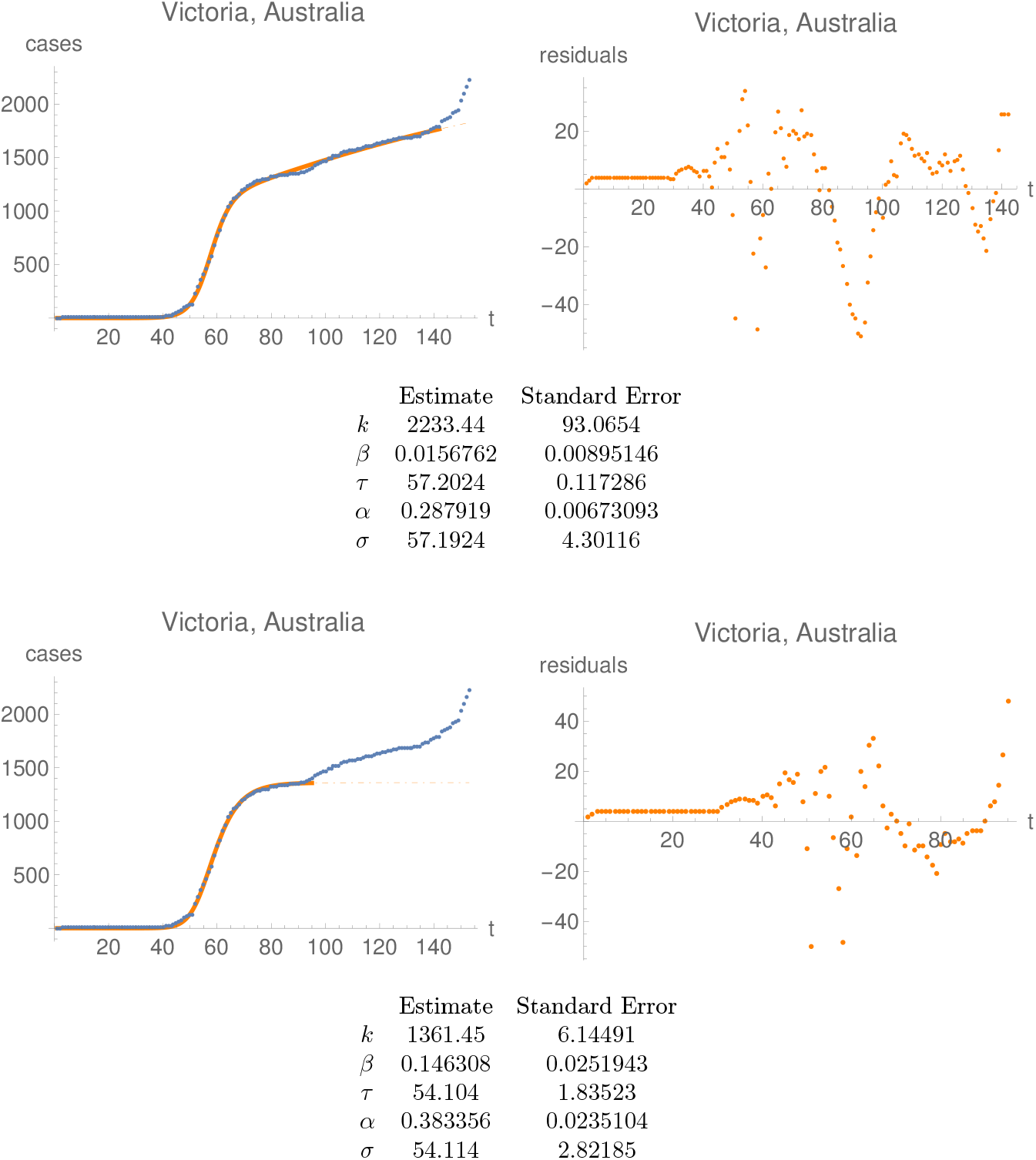
Plots of the time series and the best fit (left) before June 30, 2020, as well as the residuals of the fit (right) for the confirmed-cases time-series with smallest smallest *β*, namely the State of Victoria in Australia. Note the different scales for all plots. The data from the first plot for Victoria have been used for the analysis described in the main body of the paper. The second plot for Victoria has been fine-tuned by hand for a first phase which stops around day 82 (May 3), with optically more satisfactory residuals but with a worse “goodness parameter” (19.3% for the first plot and 24.3% for the second). Note that *τ* and *σ* are very close to each other for both plots, and we have found such fits unstable and therefore unreliable.

For the death time-series the lower bounds on the corresponding parameters are rather similar:

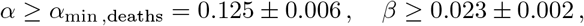

attained on the time series of DuPage, Illinois for *α* and Snohomish, Washington for *β*. This is illustrated in Figure 1_4, with the fits for DuPage and Snohomish in Figure 1_5.

**Figure 1.4:**
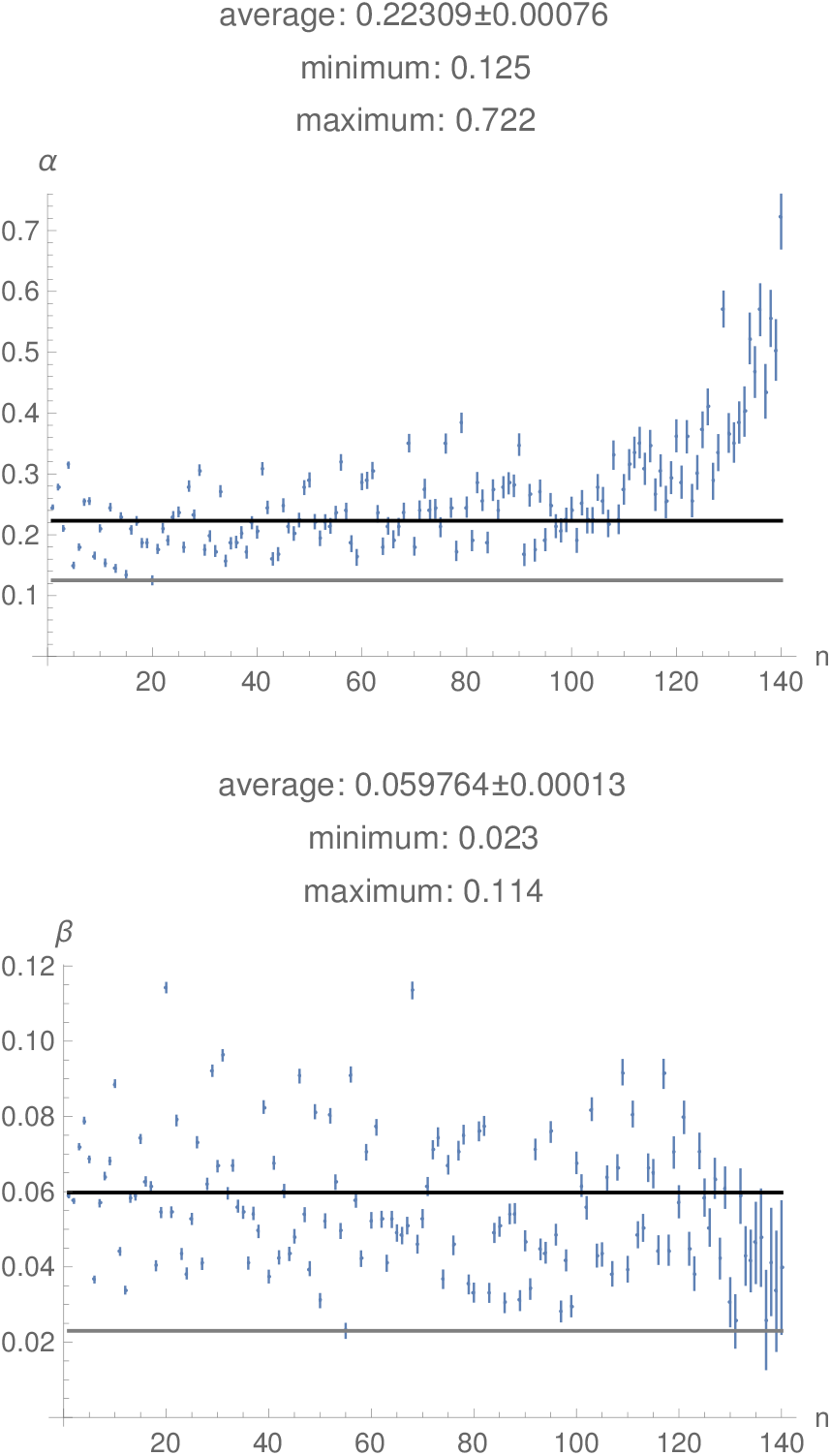
The values of the parameters *α* and *β* for the deaths time-series, ordered by increasing standard deviation. The black line is the mean and the grey line is the minimum The averages indicated here are weighted with the standard deviation, while the ones indicated in the text are the straightforward ones.

**Figure 1.5:**
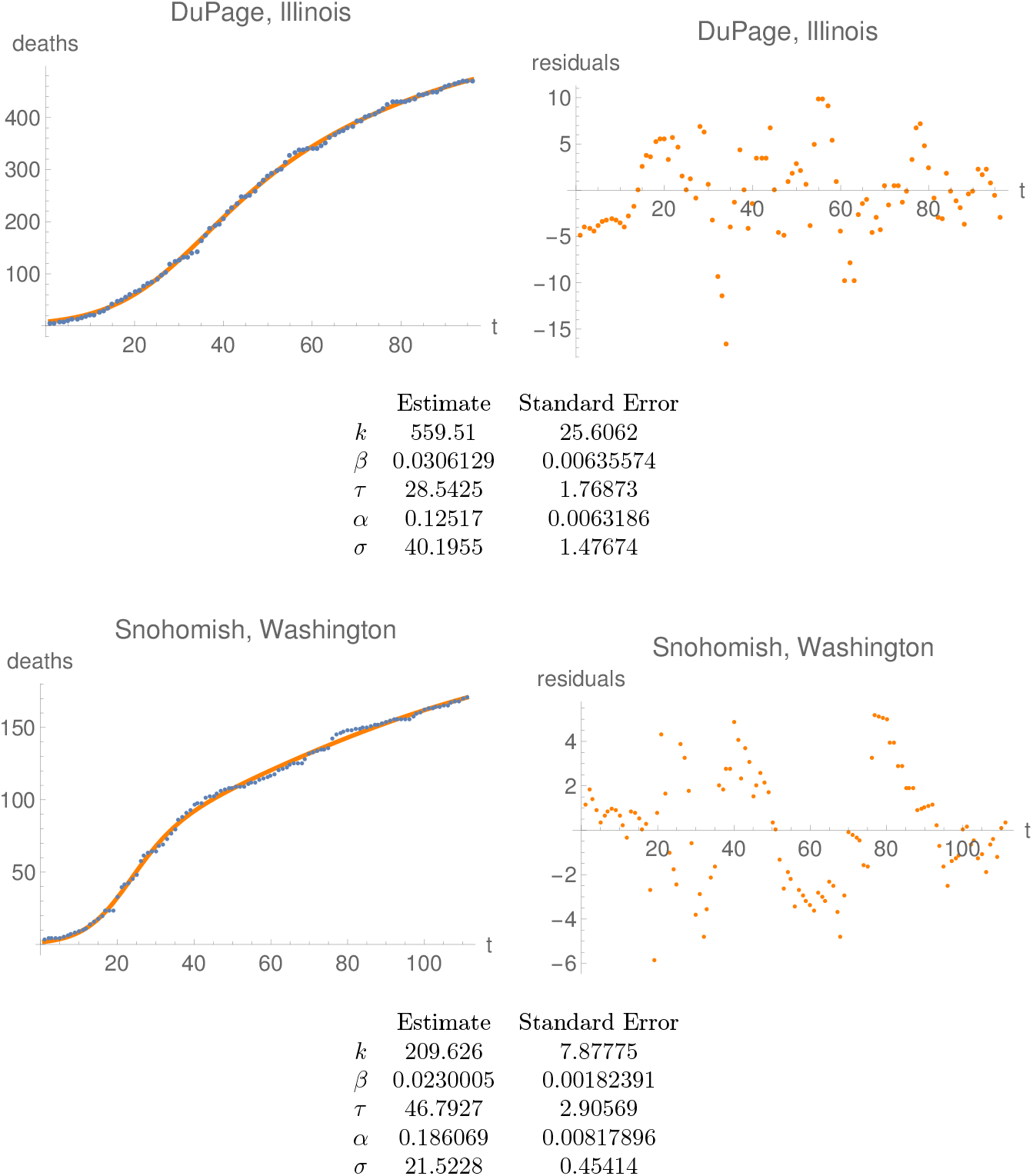
Plots of the time series and the best fit (left) before June 30, 2020, as well as the residuals of the fit (right) for the deaths time-series with smallest *α*, namely DuPage, Illinois, and smallest *β*, i.e. Snohomish, Washington. Note the different scales for all plots. Both time series end on June 30, and start at the day at which more than one death has been registered.

The parameter *α* is directly related to the *initial doubling time* of the epidemic, which will be denoted by *T*_2_. For data sets for which the approximation (1.2) holds we obtain a common bound on the doubling time:

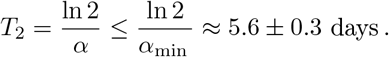

Now, for some of our fits the approximation (1.2) breaks down, but the value of the doubling time *T*_2_ can be found numerically using the explicit form of *I* when the fitting parameters are used. This shifts the threshold (1.6) to the bound

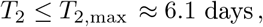

attained for the time series of Cumberland, New Jersey, already seen in Figure 1_2.

Next, for all fits (to be found in supplementary materials) we find

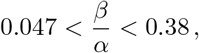

with the lower bound attained on Mason, Washington, and the upper bound on Ireland (both seen in Figure 1_6). Note that the upper bound stays well clear of the 0.5 limit imposed by our fitting function.

**Figure 1.6:**
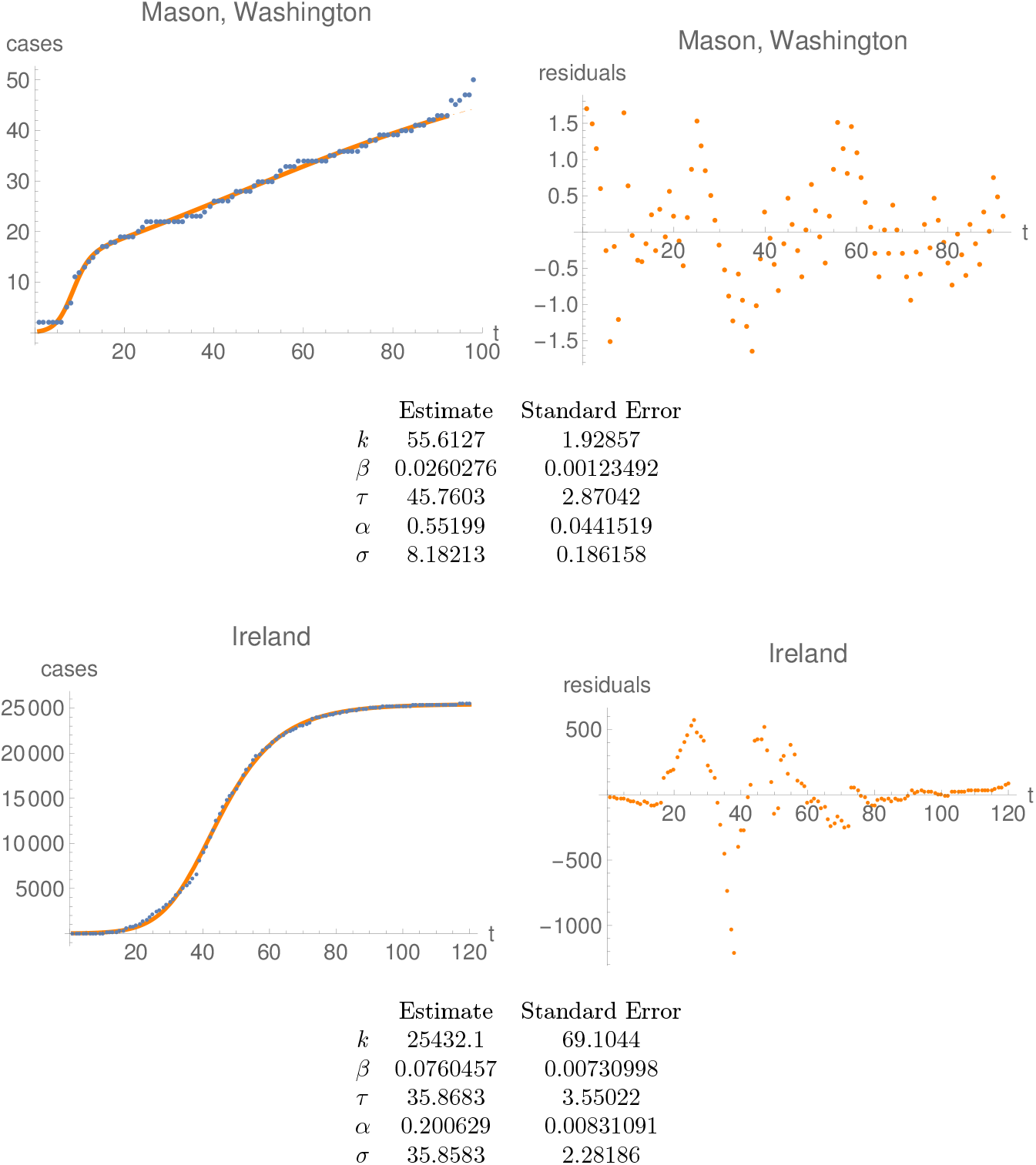
Plots of the time series and the best fit (left) before June 30, 2020, as well as the residuals of the fit (right) for the confirmed-cases time-series with smallest ratio *β/α* (for Mason, Washington) and largest ratio *β/α* (for Ireland). Note the different scales for all plots.

The average value of *β* for all satisfactory fits is

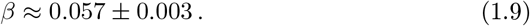

The corresponding average value for the death time-series is the same within standard deviation:

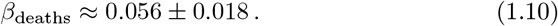

This translates to an *average time scale of twelve days for the halving of the number of new cases, or deaths, during the extinction period* of the first phase of the epidemic.

The question arises about the origin of the lower bound on *α*. Assuming that the number of confirmed cases *during the first week or so of the epidemic* is proportional to the number of infectious individuals and that the approximation (1.2) holds, we show in Appendix A that a precise lower bound *α* ≥ *α_min_* determines a lower bound on the number of days, say *k*_0_, during which an infected individual is infectious.

Supposing we had access to all possible curves for the epidemic, the threshold number *α_min_* would then be determined exactly from these curves. Now, we only have a set of 469 curves. Given that this is quite a large number, it appears reasonable to assume that the minimal value of *α* which we see in our fits is approximately equal to the threshold value *α_min_*. This, together with (1.4), leads to

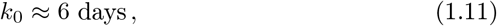

consistently with data obtained by completely different methods in [3, 4].

It should be pointed out that (1.11) applies to asymptomatic infected individuals, since the number of these is much larger than the number of symptomatic ones.

As further explained in the Appendix, for time series which are well approximated at the beginning by an exponential, the initial propagation number is determined by the equation

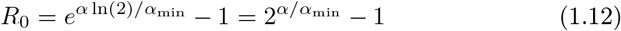

(see Equation (A.3)). This, together with (1.11), allows us to determine *R*_0_ for each time-series with a satisfactory fit.

In what follows we only consider the 145 fits for which the values of *R*_0_ have a standard deviation less than 10%. The histogram of the resulting distribution of *R*_0_ can be found in Figure 1_7. We find

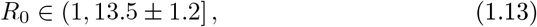

with an average

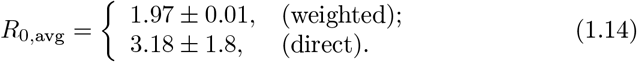

**Figure 1.7:**
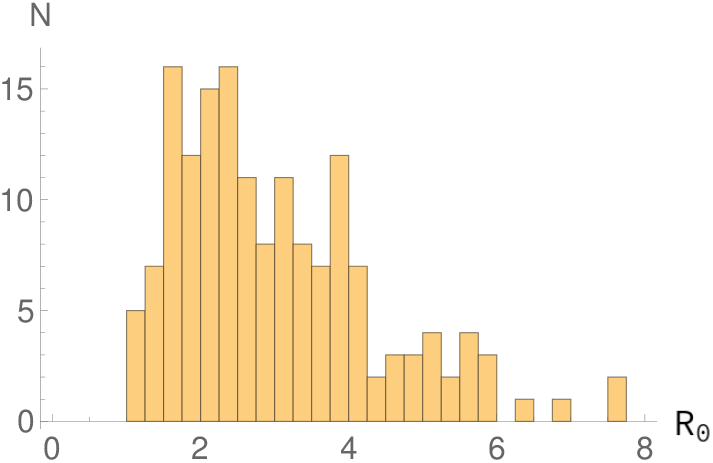
Histogram of those values of *R*_0_ which have a standard deviation less than 10% truncated at 8; see the Table B.1, Appendix B for the three missing fits beyond 8.

The weighted average is calculated by weighing with the standard deviation of *R*_0_ of the individual time-series, while the direct average is the obvious one.

The minimum value *R*_0_ =1 is attained for the county Cumberland in New Jersey, and the maximum value *R*_0_ = 13.5 ± 1.2 in (1.13) is attained for the county of Stearns in Minnesota, with fits shown in Figure 1_8 The values of *R*_0_ so determined are provided in Table 1.1 for a sample of countries. A table presenting the values of *R*_0_ for all satisfactory fits with relative error of *R*_0_ less than 10% can be found in Appendix B.

**Figure 1.8:**
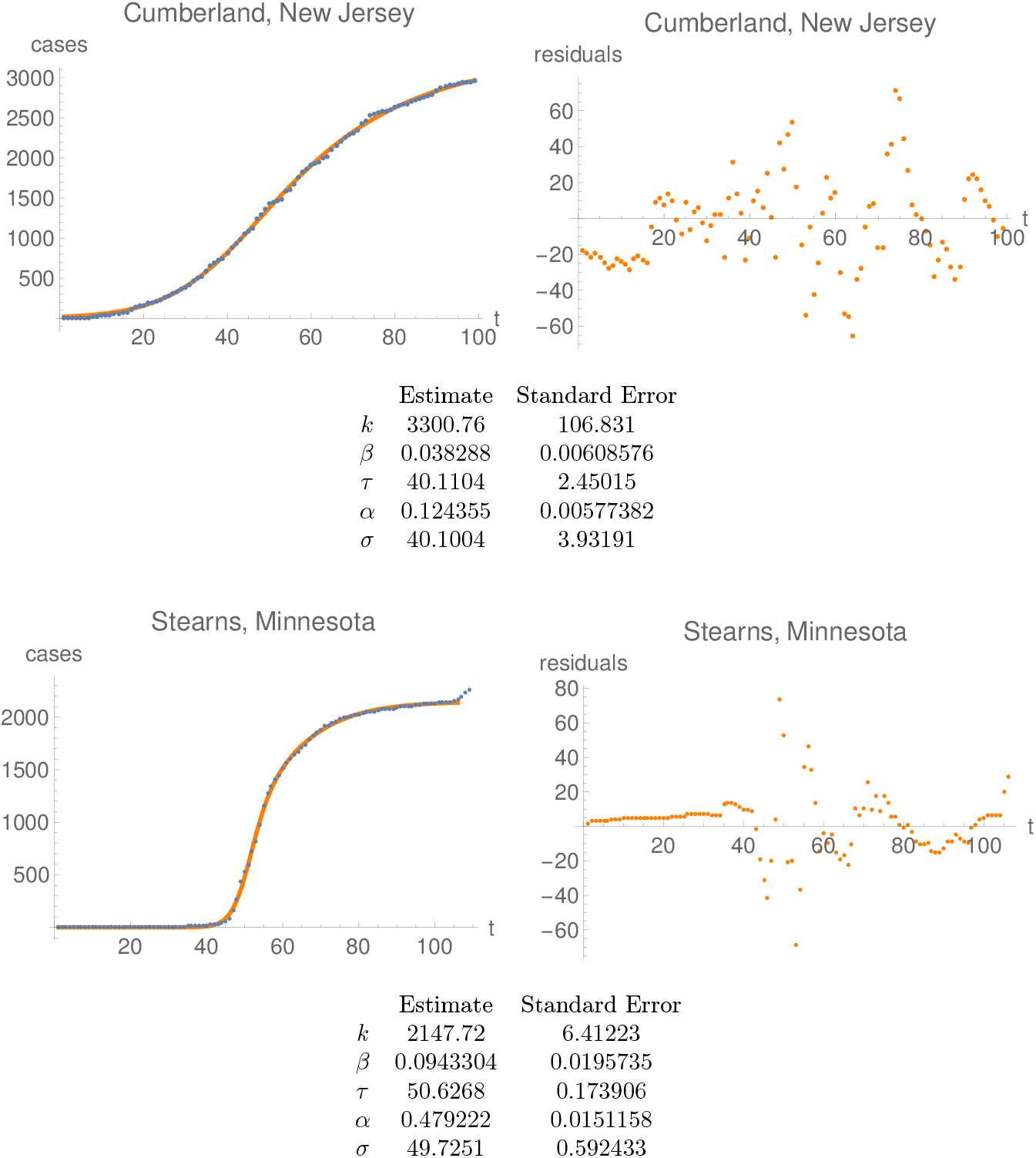
Plots of the time series and the best fit (left) before June 30, 2020, as well as the residuals of the fit (right) for the confirmed-cases time-series with the smallest (Cumberland, New Jersey) and largest (Stearns, Minnesota) value of *R*_0_. Note the different scales for all plots.

**Table1.1:** A sample of values of the initial rate of propagation *R*_0_, as determined using the double-logistic function. Strictly speaking, the numbers given are lower bounds for *R*_0_, but we expect them to be close to the correct values. The fits for the extreme members of the table, namely Austria and the US, can be found in Figure 1_9.

| | Country | $R_0$ | | Country | $R_0$ | | Country | $R_0$ |
| --- | --- | --- | --- | --- | --- | --- | --- | --- |
| 1 | Austria | $5.02 \pm 0.24$ | 9 | Germany | $2.67 \pm 0.06$ | 17 | Netherlands | $4.8 \pm 0.5$ |
| 2 | Belgium | $1.85 \pm 0.07$ | 10 | Greece | $1.92 \pm 0.09$ | 18 | Poland | $1.86 \pm 0.07$ |
| 3 | Croatia | $5.5 \pm 0.5$ | 11 | Hungary | $1.51 \pm 0.07$ | 19 | Romania | $2.63 \pm 0.11$ |
| 4 | Cyprus | $3.00 \pm 0.25$ | 12 | Ireland | $2.06 \pm 0.14$ | 20 | Spain | $2.56 \pm 0.11$ |
| 5 | Czechia | $2.71 \pm 0.13$ | 13 | Italy | $3.07 \pm 0.07$ | 21 | Sweden | $1.24 \pm 0.06$ |
| 6 | Denmark | $1.29 \pm 0.06$ | 14 | Latvia | $3.94 \pm 0.21$ | 22 | United Kingdom | $2.72 \pm 0.11$ |
| 7 | Estonia | $2.27 \pm 0.20$ | 15 | Lithuania | $2.13 \pm 0.16$ | 23 | US | $3.38 \pm 0.09$ |
| 8 | Finland | $2.12 \pm 0.14$ | 16 | Luxembourg | $5.9 \pm 0.4$ | | | |

## 2 The analysis

We analysed the time series for confirmed cases, confirmed US cases, deaths and US deaths as available on the John Hopkins University server on July 1, 2020. After ignoring all time-series which have less than two cases on June 30, as well as the US time-series which appear as “Unassigned” and “Out of …”, we were left with 3149 confirmed-cases time-series and 1647 deaths time-series.

These time series include whole countries, but also time series for e.g. French Domaines d’Outre Mer-Territoires d’Outre Mer, or for each Australian State, or for each Chinese Province, or for US counties.

**Figure 1.9:**
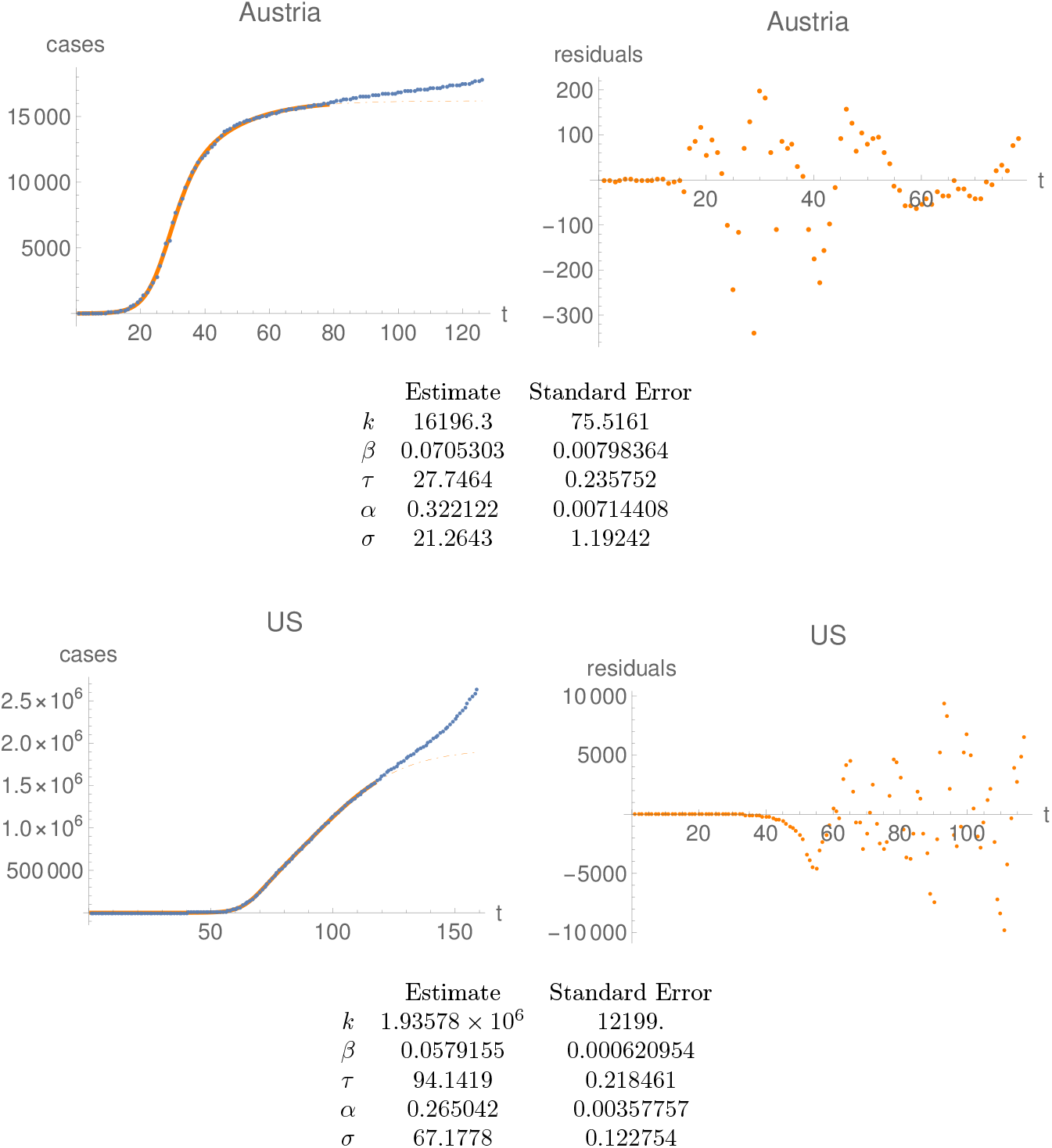
Plots of the time series and the best fit (left) before June 30, 2020, as well as the residuals of the fit (right) for the confirmed-cases time-series for Austria and for the US. Note the different scales for all plots.

For each such time-series we attempted to determine the day, say *T_f_*, and the parameters (*α, β, k, τ, σ*) so that the curve (1.1) fits optimally the data up to day *T_f_*, using the `NonlinearModelFit` procedure in MATHEMATICA. Finding a fit sometimes requires indicating parameters which are used as starting values for the fitting procedure, with fits depending upon the starting parameters. To take this into account we generated, for each time-series, a hundred random values of starting parameters, and from the collection of resulting fits we chose the “best fit”. This “best fit” was determined by adding the values of the largest relative residual (defined as the ratio of the largest residual to the number of cases at the end of the time series) to the relative standard deviations of all five parameters of the fit, and choosing the fit for which the resulting number was smallest. We carried-out this analysis for time series truncated to April 16, then to April 17, etc., until June 30, and the day of the *end of the first phase*, say *T_f_*, was determined by choosing the time series with the best fit on day *T_f_*. All the satisfactory fits found had a date *T_f_* later than April 27 (with April 28 attained by the time series for Sichuan, China, seen in Figure 2_1.)

**Figure 2.1:**
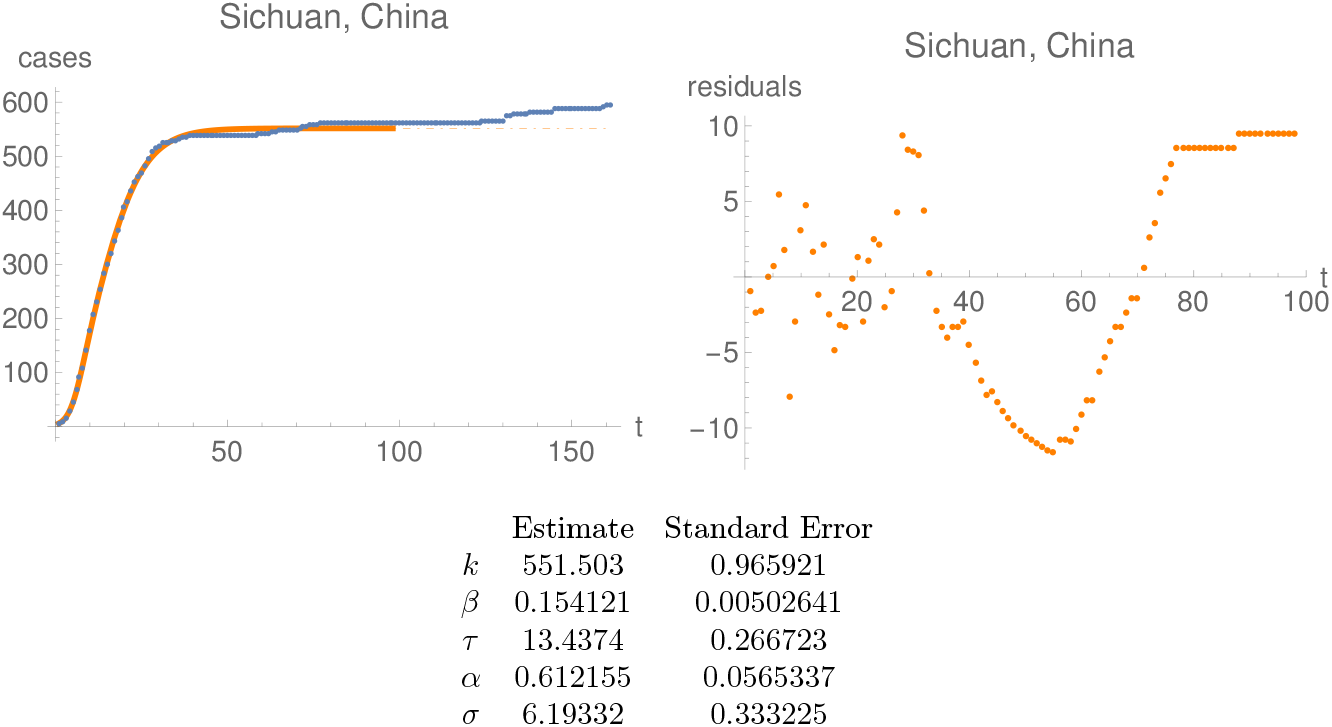
Plots of the time series and the best fit (left) before June 30, 2020, as well as the residuals of the fit (right) for the confirmed-cases time-series of Sichuan, China, which has the earliest date of the end of the first phase of the pandemic. Note the different scales for all plots.

While the above results in optically satisfactory fits in most cases, some of the resulting fits have parameters with large standard deviations. To eliminate those we only kept the fits where the absolute value of the largest relative residual divided by the last value of the time series was smaller than 10%, and where the relative standard deviations of each parameter were smaller than10%. All this led to 469 satisfactory fits for the confirmed-cases time-series, and 140 satisfactory fits for the deaths time-series.

In Figure1.1 we show the resulting values of the parameters *α* and *β* together with their standard errors, ordered by increasing length of confidence interval, for the confirmed-cases time-series. The figure shows clearly the lower bound of *α* of (1.4). Figure 2_2 shows the histograms of the values of *α* and *β*.

It follows from (1.2) that the larger the value of *α*, the faster the initial growth of the epidemic. So the two time-series with smallest value of *α*, namely Cumberland, New Jersey (see Figure 1_8) and San Juan, New Mexico (Figure 2_3) are the ones with the slowest initial growth of the epidemic. Likewise the two time-series with largest values of *α*, namely South Korea and Stearns, Minnesota, are the ones with fastest initial growth of the epidemic. The fits for these time series are shown in Figures 1.8 and 2.4.

**Figure 2.2:**
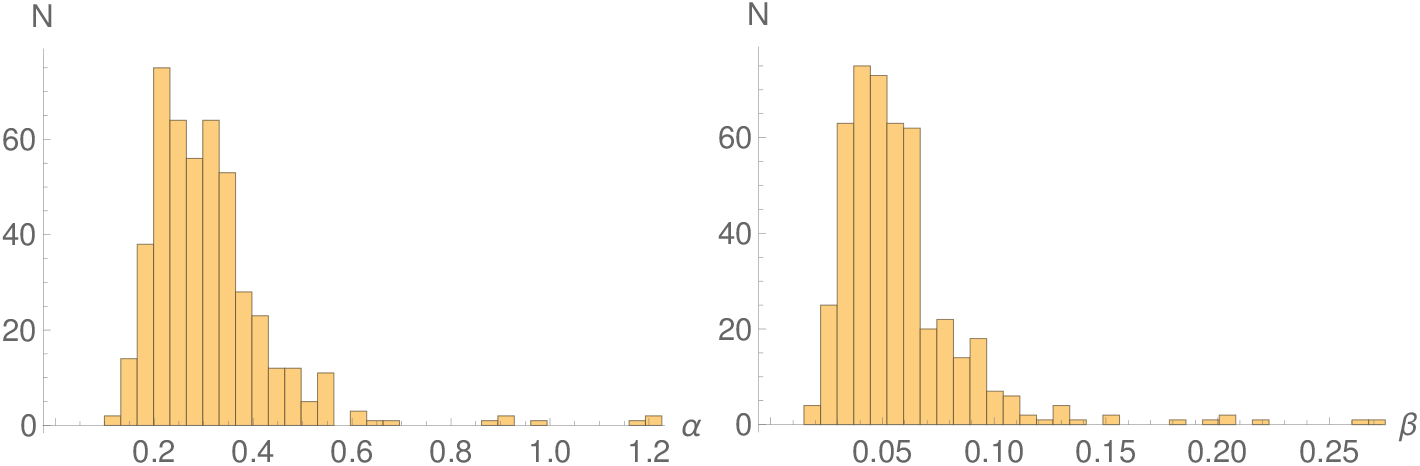
Histograms of the values of *α* (left) and *β* (right) for the confirmed-cases time-series.

The last data point on all plots is June 30, 2020.

We note that we only show in this paper the fits for the outliers of our analysis, which are not necessarily very good fits, and we hope that the reader will not draw the conclusion that no good fits can be obtained to the curve (1.1). In the supplementary material the reader will find the complete set of our fits, which hopefully should dispel such an impression. It should also be clear, by inspection of the fits presented here or in the supplementary materials, that the fits can be fine-tuned for individual time-series by removing outliers and other data manipulation, but we have not attempted to do this.

We have repeated the above analysis for the deaths time-series from the JHU server. Figure 1_4 illustrates the lower bound *α* ≥ 0.125 for these time series.

**Figure 2.3:**
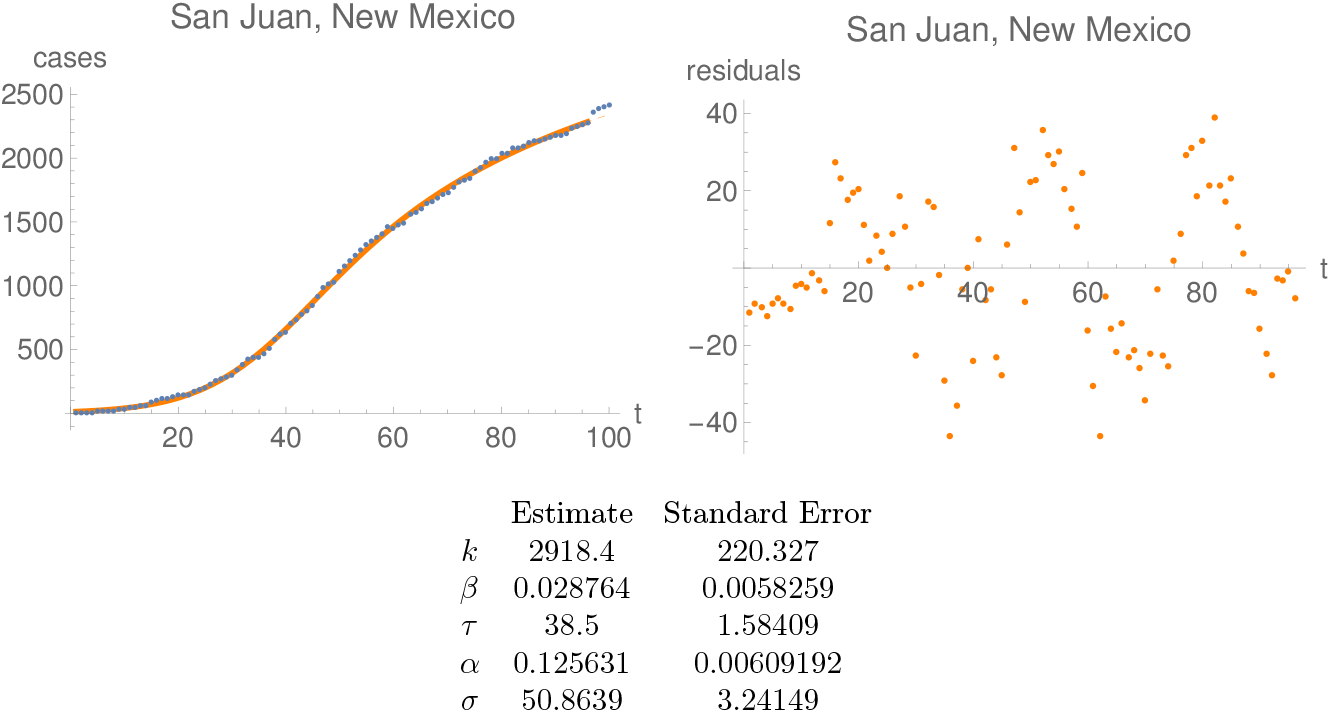
Plots of the time series and the best fit (left) before June 30, 2020, as well as the residuals of the fit (right) for the confirmed-cases time-series of San Juan, New Mexico, with the second smallest value of *α*. Note the different scales for the plots.

The relevant histograms can be found in Figure 2_6. The lower bound for *α* is attained on the time series of DuPage, Illinois (*α* =0.125 ±0.006) with the next smallest value for Riverside, California (*α* =0.135 ± 0.006). The two deaths time-series recording the fastest initial growth rate are Westchester, New York (*α* =0.57±0.03), and Nassau, New York (*α* =0.72±0.05). The four associated fits can be seen in Figures 1.5, 2.7 and 2.8.

In Figure 2.5 we show the values of *α* and *β* for all fits for which the parameter errors are less than 100%; the values of the parameters for such fits, when not in our previous list filtered to 10%, should be interpreted with care. The plot shows that our lower bounds (1.4) are satisfied by an overwhelming majority of fits, including the poor ones.

## 3 Conclusions

We used the double-logistic curve of [1] to obtain quantitative information on the dynamics of the Covid-19 epidemics up to June 30, 2020. This curve approximates surprisingly well most of the time series until a date which we interpret as the end of the first phase of the epidemic, even when the fits are unsatisfactory from a mathematical point of view. In many cases the fact that the errors of the fitting parameters are large can be explained by a short time-series, or changes in the reporting procedures during the epidemic, or appearances of new clusters related to changes in confinement measures.

We have shown existence of a lower rate of the initial growth of the epidemics. We suggest that this lower bound can be used to determine the time during which an infected asymptomatic individual is infectious. This then allows one to determine the initial propagation number *R*_0_ for time series with satisfactory fits to our curve. We have observed that the average extinction rate of the epidemics is about 12 days.

**Figure 2.4:**
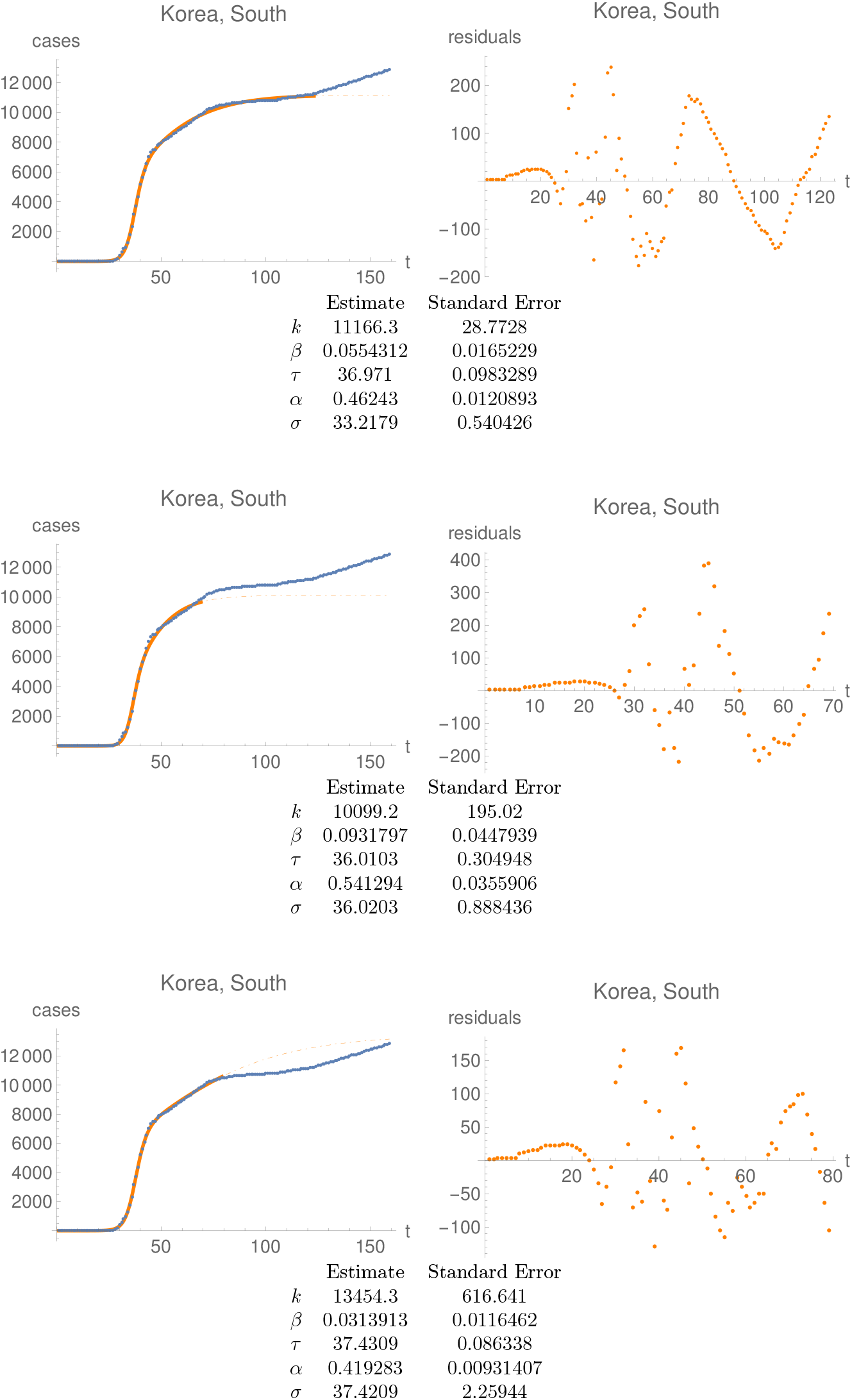
Data points, the best fit before June 30, 2020, and its residuals for South Korea. The first fit, automatically chosen by our algorithm and used for the analysis in the body of the paper, indicates existence of two phases, with the second one starting on May 26. The second fit has been fine-tuned by hand to find a second phase starting around day 50 of the epidemics; in this case the fit ends on April 1. The third fit ends on April 11 and has been found by the computer as one with the best goodness parameter for fits ending around the beginning of April. These two fits illustrate again that fits with very similar *τ* and *σ* are unstable. One can wonder in any case whether any systematic studies based on our curve (1.1) apply to the time series of South Korea.

**Figure 2.5:**
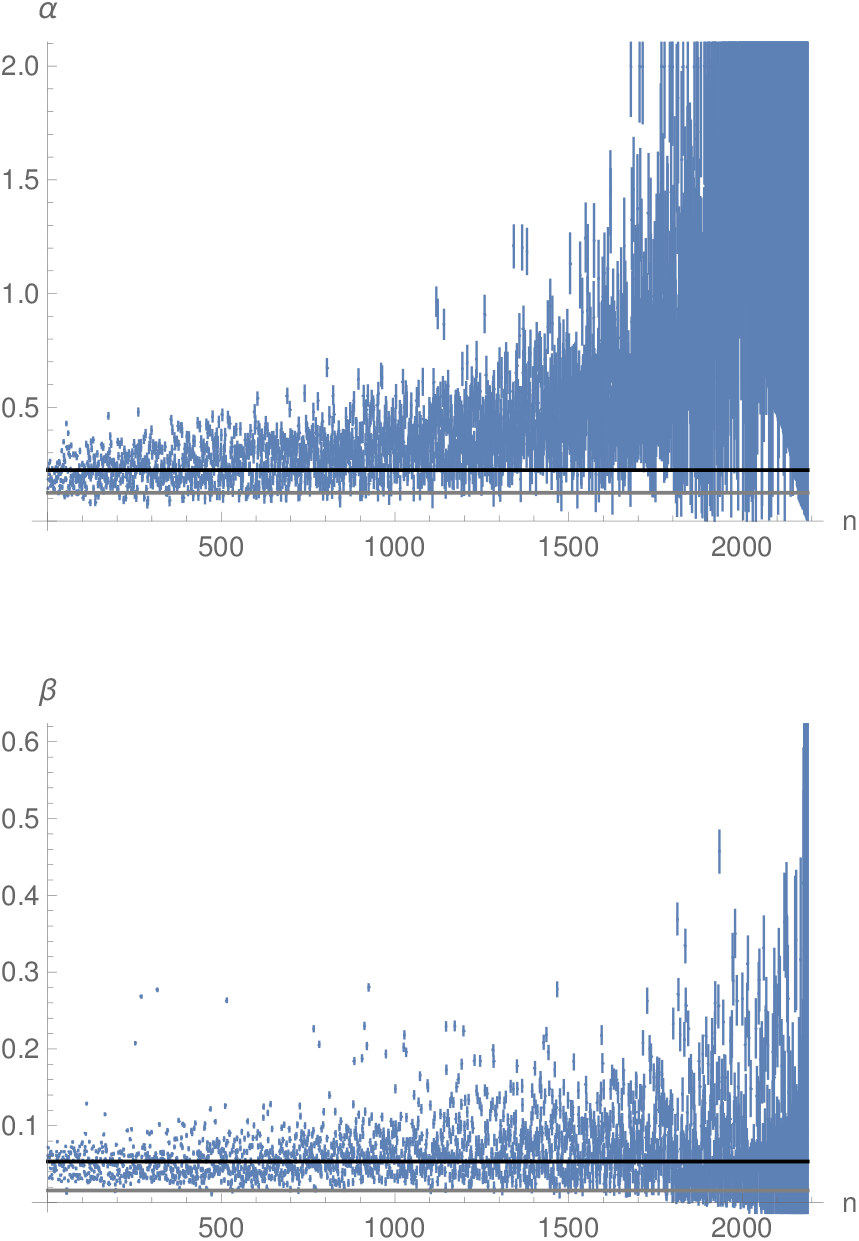
The values of the parameters *α* and *β* for the confirmed-cases time-series, ordered by increasing standard deviation, after rejecting fits with standard deviations larger than 100% on some parameter. The black line is the mean weighted by standard deviation and the grey lines are the value of *α_min_* and *β_min_* obtained from the analysis based on the fits with standard deviations for all parameters less than 10%. The cut-off at 2 of *α* in the first plot is an artifact of our fitting method.

**Figure 2.6:**
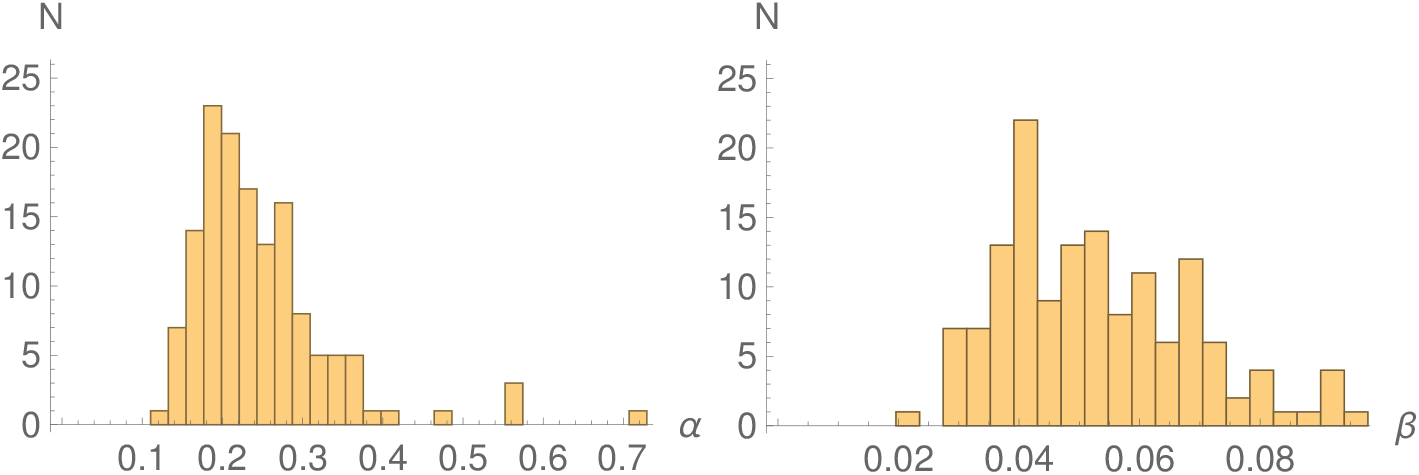
Histograms of the values of *α* and *β* for the deaths time-series.

**Figure 2.7:**
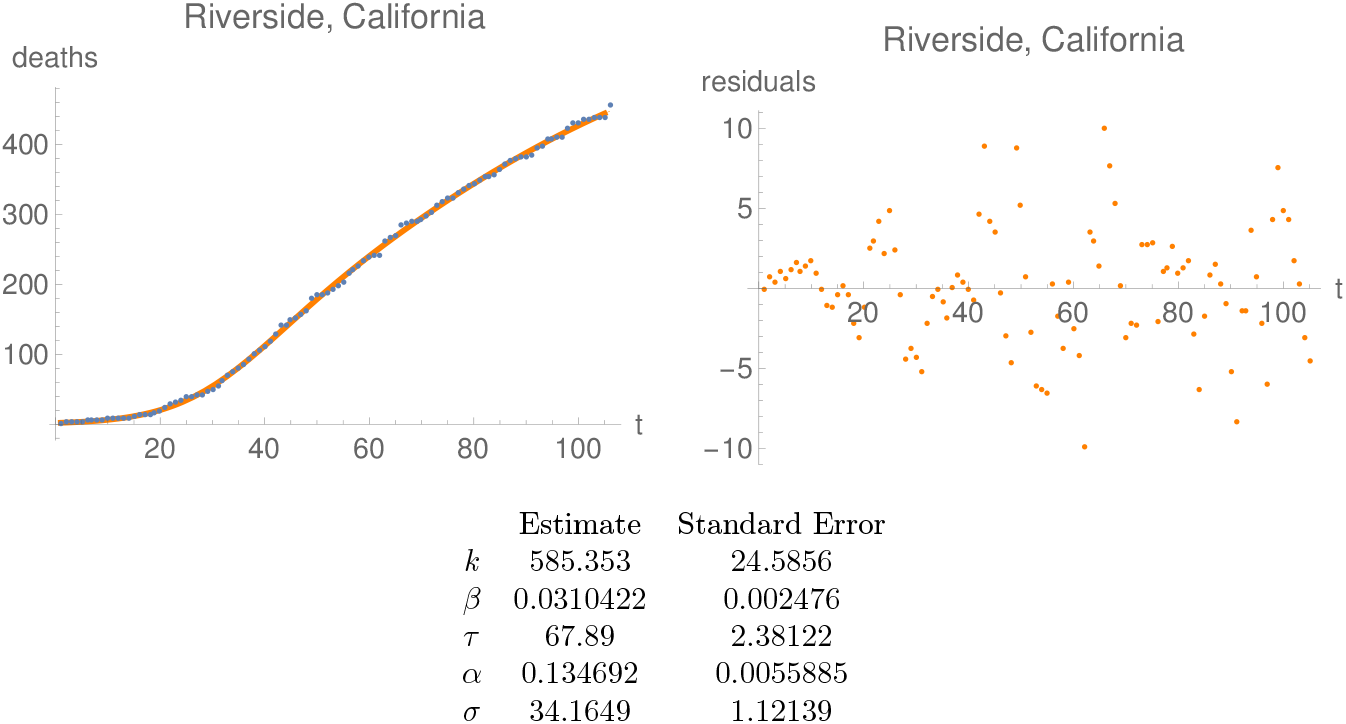
Plots of the time series for the best fits before June 30, 2020, and the fit (left), as well as the residuals (right) of the fit for the deaths time-series with the second smallest value of *α*, namely Riverside, California.

It should be admitted that our analysis gives unambiguous results only for these time series where a mathematically satisfactory fit to the time series has been found, and might be biased by the requirement that the extinction rate is smaller than half of the initial growth rate. It is therefore justified to question the universal validity of our findings, even more so because of the ad-hoc character of the curve (1.1).

### A The growth rate

Consider an interval of data 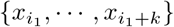 describing the number of *infectious* individuals in *k* +1 consecutive days. (We expect that the numbers *x_i_* are proportional to the number of confirmed cases at the beginning of the epidemic. This is relevant for our interpretation in the main body of this paper, but it is irrelevant for the considerations in this section where *x_i_* denotes, by definition, the number of infectious individuals.) Suppose that the data are well approximated by the exponential curve

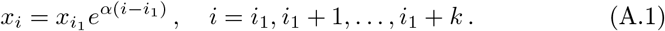

Let *k*_0_ denote the number of days during which an individual is infectious. We suppose that the probability of infecting someone is constant during this period. At day *i*_1_ + *k* with *k* ≤ *k*_0_, thus after *k* days, each individual which is infections at day *i1* will lead to *e^αk^* infected individuals, hence will have infected

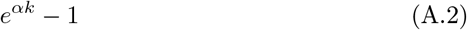

individuals.

After *k*_0_ days the individual is removed from the set of infectious individuals, hence the remaining number of infectious individuals equals

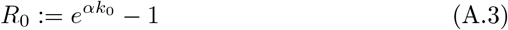

**Figure 2.8:**
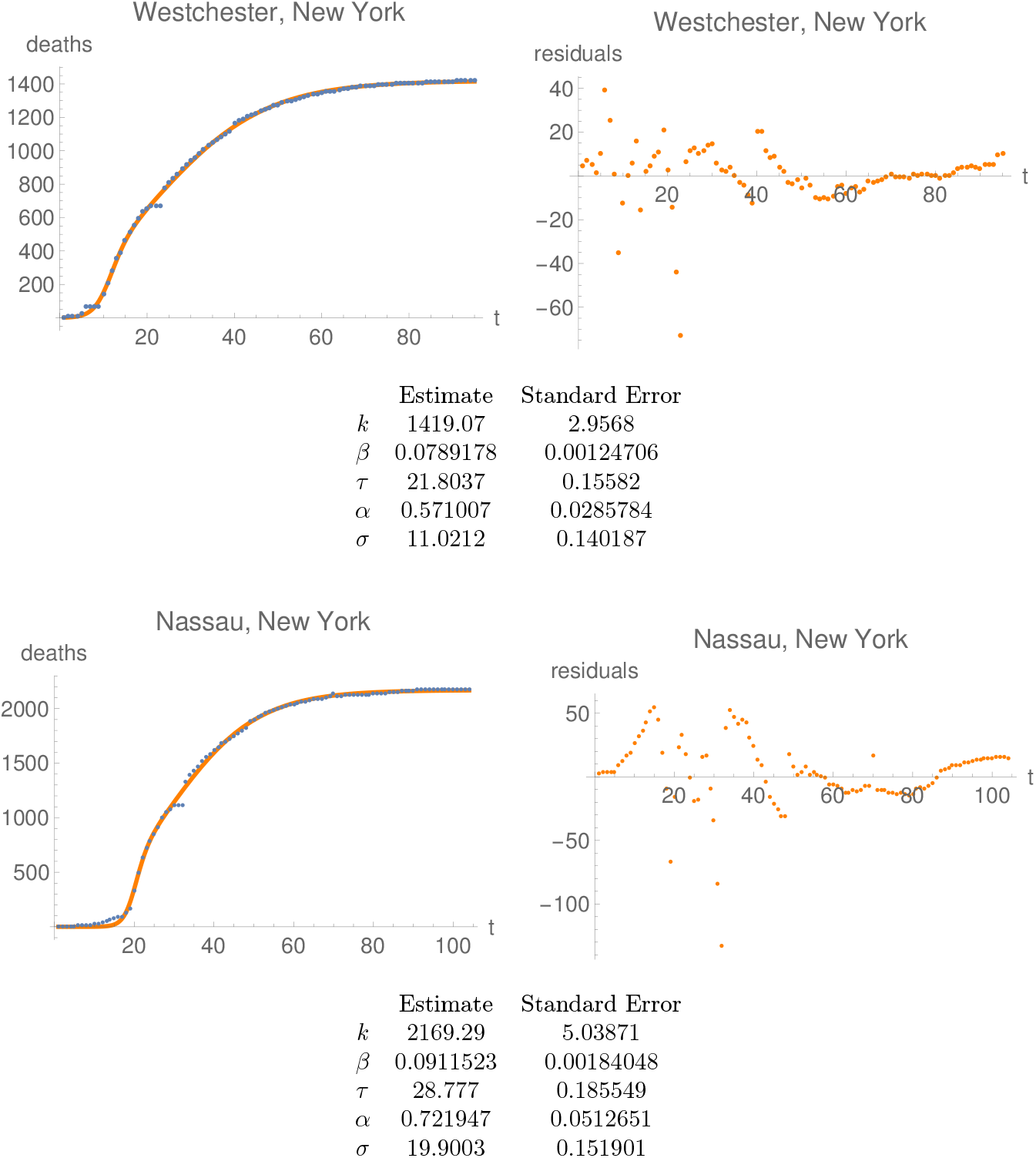
Plots of the time series for the best fits before June 30, 2020, and the fit (left), as well as the residuals (right) of the fit for the deaths time-series with largest values of *α* (Westchester and Nassau Counties, both New York State). One could fine-tune both fits by smoothing out the jumps, which drive the fit down and therefore the residuals up.

It follows that the law (A.1) persists when 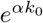 is much larger than one. On the other hand, (A.1) will break down when 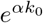 is of order one.

Within this model, an epidemic dies out if the number of infectious individuals after *k*_0_ days is smaller than 1:

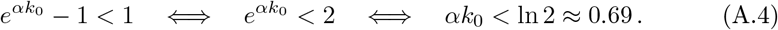

Thus, an epidemic time-series for which

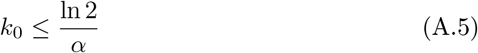

never follows the exponential law (A.1) for more than *k*_0_ days.

Equivalently, *no epidemic with the exponential law* (A.1) *will be observed* with a parameter *α* such that

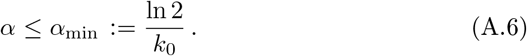

We conclude that the time *k*_0_ during which an individual is infectious can be determined by measuring the threshold number *αmin*.

Note that the equality case in (A.4) corresponds to the doubling time of the epidemic. Hence, in this simple model, *the time during which an individual is infectious is smaller than or equal the smallest observed doubling time of the epidemic*.

### B *R*_0_ for selected time-series

**Table B.1:** The initial rate of propagation *R*_0_ for time series with a relative standard deviation smaller than 10%, ordered according to the value of *R*_0_. Strictly speaking, the numbers given are lower bounds for *R*_0_, but we expect them to be close to the correct values.

| | Country | $R_0$ | | Country | $R_0$ | | Country | $R_0$ |
| --- | --- | --- | --- | --- | --- | --- | --- | --- |
| 1 | Cumberland, New Jersey | $1.00 \pm 0.06$ | 56 | Snohomish, Washington | $2.26 \pm 0.09$ | 111 | Collin, Texas | $3.52 \pm 0.29$ |
| 2 | San Juan, New Mexico | $1.01 \pm 0.07$ | 57 | Estonia | $2.27 \pm 0.20$ | 112 | Marshall, Iowa | $3.53 \pm 0.29$ |
| 3 | Mexico | $1.15 \pm 0.06$ | 58 | Saginaw, Michigan | $2.31 \pm 0.20$ | 113 | Jefferson, Colorado | $3.55 \pm 0.29$ |
| 4 | Russia | $1.240 \pm 0.028$ | 59 | Fairfield, Connecticut | $2.32 \pm 0.19$ | 114 | Somerset, New Jersey | $3.57 \pm 0.35$ |
| 5 | Sweden | $1.24 \pm 0.06$ | 60 | Lake, Indiana | $2.32 \pm 0.15$ | 115 | Osceola, Florida | $3.57 \pm 0.26$ |
| 6 | Denmark | $1.29 \pm 0.06$ | 61 | Calhoun, Michigan | $2.32 \pm 0.22$ | 116 | Sandoval, New Mexico | $3.69 \pm 0.28$ |
| 7 | Harford, Maryland | $1.29 \pm 0.08$ | 62 | Mercer, New Jersey | $2.34 \pm 0.18$ | 117 | Delaware, Pennsylvania | $3.78 \pm 0.30$ |
| 8 | Kent, Michigan | $1.29 \pm 0.08$ | 63 | Anne Arundel, Maryland | $2.34 \pm 0.18$ | 118 | Cuyahoga, Ohio | $3.79 \pm 0.31$ |
| 9 | Los Angeles, California | $1.31 \pm 0.09$ | 64 | San Mateo, California | $2.35 \pm 0.21$ | 119 | Brown, Wisconsin | $3.8 \pm 0.4$ |
| 10 | Indonesia | $1.33 \pm 0.08$ | 65 | Rockingham, Virginia | $2.38 \pm 0.19$ | 120 | Berks, Pennsylvania | $3.79 \pm 0.22$ |
| 11 | St. Clair, Illinois | $1.44 \pm 0.13$ | 66 | Pima, Arizona | $2.39 \pm 0.16$ | 121 | St. Louis, Missouri | $3.80 \pm 0.28$ |
| 12 | Cuba | $1.46 \pm 0.06$ | 67 | Washington, Oregon | $2.39 \pm 0.12$ | 122 | Berkshire, Massachusetts | $3.83 \pm 0.27$ |
| 13 | Hungary | $1.51 \pm 0.07$ | 68 | Singapore | $2.42 \pm 0.14$ | 123 | Middlesex, Connecticut | $3.9 \pm 0.4$ |
| 14 | Rock Island, Illinois | $1.56 \pm 0.14$ | 69 | Montgomery, Pennsylvania | $2.44 \pm 0.11$ | 124 | Middlesex, New Jersey | $3.9 \pm 0.4$ |
| 15 | Guinea | $1.59 \pm 0.11$ | 70 | Salt Lake, Utah | $2.46 \pm 0.18$ | 125 | Lebanon, Pennsylvania | $3.9 \pm 0.4$ |
| 16 | Geauga, Ohio | $1.59 \pm 0.13$ | 71 | Norway | $2.46 \pm 0.12$ | 126 | Latvia | $3.94 \pm 0.21$ |
| 17 | Serbia | $1.60 \pm 0.07$ | 72 | Pierce, Washington | $2.53 \pm 0.15$ | 127 | Victoria, Australia | $3.98 \pm 0.19$ |
| 18 | King, Washington | $1.61 \pm 0.07$ | 73 | Spain | $2.56 \pm 0.11$ | 128 | Orange, New York | $4.0 \pm 0.4$ |
| 19 | Chester, Pennsylvania | $1.64 \pm 0.10$ | 74 | Burkina Faso | $2.60 \pm 0.22$ | 129 | Summit, Utah | $4.05 \pm 0.34$ |
| 20 | Essex, Massachusetts | $1.67 \pm 0.08$ | 75 | Bartow, Georgia | $2.60 \pm 0.24$ | 130 | Tallapoosa, Alabama | $4.1 \pm 0.4$ |
| 21 | Howard, Maryland | $1.67 \pm 0.17$ | 76 | Camden, New Jersey | $2.62 \pm 0.14$ | 131 | Allegheny, Pennsylvania | $4.11 \pm 0.24$ |
| 22 | Ukraine | $1.68 \pm 0.08$ | 77 | Romania | $2.63 \pm 0.11$ | 132 | Thailand | $4.13 \pm 0.23$ |
| 23 | Cherokee, Georgia | $1.69 \pm 0.14$ | 78 | Nassau, New York | $2.64 \pm 0.09$ | 133 | New York, New York | $4.19 \pm 0.32$ |
| 24 | Suffolk, Massachusetts | $1.69 \pm 0.08$ | 79 | Germany | $2.67 \pm 0.06$ | 134 | Black Hawk, Iowa | $4.22 \pm 0.21$ |
| 25 | Norfolk, Massachusetts | $1.70 \pm 0.08$ | 80 | Czechia | $2.71 \pm 0.13$ | 135 | Kandiyohi, Minnesota | $4.2 \pm 0.4$ |
| 26 | Middlesex, Massachusetts | $1.70 \pm 0.05$ | 81 | Prince George's, Maryland | $2.71 \pm 0.27$ | 136 | Livingston, Michigan | $4.3 \pm 0.4$ |
| 27 | India | $1.70 \pm 0.09$ | 82 | United Kingdom | $2.72 \pm 0.11$ | 137 | Philippines | $4.49 \pm 0.30$ |
| 28 | Dauphin, Pennsylvania | $1.74 \pm 0.12$ | 83 | Panama | $2.76 \pm 0.22$ | 138 | Dutchess, New York | $4.57 \pm 0.28$ |
| 29 | Ontario, Canada | $1.77 \pm 0.07$ | 84 | Bangladesh | $2.78 \pm 0.20$ | 139 | Wayne, Michigan | $4.60 \pm 0.20$ |
| 30 | San Francisco, California | $1.77 \pm 0.11$ | 85 | Denton, Texas | $2.78 \pm 0.21$ | 140 | Ocean, New Jersey | $4.7 \pm 0.4$ |
| 31 | Belarus | $1.79 \pm 0.04$ | 86 | Webb, Texas | $2.82 \pm 0.19$ | 141 | Netherlands | $4.8 \pm 0.5$ |
| 32 | Litchfield, Connecticut | $1.80 \pm 0.11$ | 87 | Palm Beach, Florida | $2.92 \pm 0.24$ | 142 | Pike, Pennsylvania | $4.8 \pm 0.5$ |
| 33 | Belgium | $1.85 \pm 0.07$ | 88 | Fulton, Georgia | $2.92 \pm 0.29$ | 143 | Genesee, Michigan | $4.88 \pm 0.34$ |
| 34 | Poland | $1.86 \pm 0.07$ | 89 | Richland, South Carolina | $2.94 \pm 0.23$ | 144 | Austria | $5.02 \pm 0.24$ |
| 35 | Cobb, Georgia | $1.86 \pm 0.14$ | 90 | Philadelphia, Pennsylvania | $2.99 \pm 0.27$ | 145 | Washtenaw, Michigan | $5.05 \pm 0.34$ |
| 36 | DeKalb, Georgia | $1.87 \pm 0.16$ | 91 | Cyprus | $3.00 \pm 0.25$ | 146 | Jefferson, Louisiana | $5.08 \pm 0.25$ |
| 37 | McKinley, New Mexico | $1.91 \pm 0.09$ | 92 | Oakland, Michigan | $3.06 \pm 0.13$ | 147 | Morris, New Jersey | $5.1 \pm 0.4$ |
| 38 | Greece | $1.92 \pm 0.09$ | 93 | Italy | $3.07 \pm 0.07$ | 148 | Miami-Dade, Florida | $5.4 \pm 0.4$ |
| 39 | Alameda, California | $1.99 \pm 0.14$ | 94 | Houston, Georgia | $3.09 \pm 0.31$ | 149 | Switzerland | $5.38 \pm 0.28$ |
| 40 | Douglas, Georgia | $2.00 \pm 0.18$ | 95 | Scott, Iowa | $3.09 \pm 0.23$ | 150 | Croatia | $5.5 \pm 0.5$ |
| 41 | Scott, Mississippi | $2.01 \pm 0.16$ | 96 | District of Columbia | $3.10 \pm 0.25$ | 151 | Essex, New Jersey | $5.60 \pm 0.35$ |
| 42 | Adams, Pennsylvania | $2.04 \pm 0.20$ | 97 | New Haven, Connecticut | $3.15 \pm 0.13$ | 152 | St. Tammany, Louisiana | $5.6 \pm 0.5$ |
| 43 | Santa Clara, California | $2.04 \pm 0.14$ | 98 | Carbon, Pennsylvania | $3.16 \pm 0.25$ | 153 | Channel Islands, United Kingdom | $5.7 \pm 0.5$ |
| 44 | Ireland | $2.06 \pm 0.14$ | 99 | De Soto, Louisiana | $3.17 \pm 0.30$ | 154 | Bergen, New Jersey | $5.9 \pm 0.5$ |
| 45 | St. Joseph, Indiana | $2.06 \pm 0.19$ | 100 | Trumbull, Ohio | $3.23 \pm 0.30$ | 155 | Luxembourg | $5.9 \pm 0.4$ |
| 46 | Dominican Republic | $2.07 \pm 0.17$ | 101 | Hampden, Massachusetts | $3.25 \pm 0.28$ | 156 | Monroe, Pennsylvania | $5.98 \pm 0.26$ |
| 47 | Ulster, New York | $2.08 \pm 0.16$ | 102 | Boone, Indiana | $3.28 \pm 0.30$ | 157 | Lafourche, Louisiana | $6.3 \pm 0.4$ |
| 48 | Finland | $2.12 \pm 0.14$ | 103 | Broward, Florida | $3.28 \pm 0.27$ | 158 | Hamilton, Indiana | $7.0 \pm 0.5$ |
| 49 | Lithuania | $2.13 \pm 0.16$ | 104 | Waukesha, Wisconsin | $3.30 \pm 0.33$ | 159 | Northampton, Pennsylvania | $7.5 \pm 0.7$ |
| 50 | Turkey | $2.16 \pm 0.06$ | 105 | Monmouth, New Jersey | $3.31 \pm 0.14$ | 160 | Lehigh, Pennsylvania | $7.61 \pm 0.35$ |
| 51 | Leake, Mississippi | $2.16 \pm 0.14$ | 106 | Macomb, Michigan | $3.31 \pm 0.17$ | 161 | Luzerne, Pennsylvania | $9.9 \pm 0.4$ |
| 52 | Moldova | $2.17 \pm 0.15$ | 107 | US | $3.38 \pm 0.09$ | 162 | Korea, South | $12.2 \pm 0.9$ |
| 53 | Crisp, Georgia | $2.18 \pm 0.17$ | 108 | Charles, Maryland | $3.40 \pm 0.30$ | 163 | Stearns, Minnesota | $13.5 \pm 1.2$ |
| 54 | York, Pennsylvania | $2.20 \pm 0.10$ | 109 | Guinea-Bissau | $3.45 \pm 0.31$ | | | |
| 55 | Washington, Iowa | $2.21 \pm 0.11$ | 110 | Suffolk, New York | $3.51 \pm 0.17$ | | | |

### C Remarks on the “double-logistic” function

Consider the function

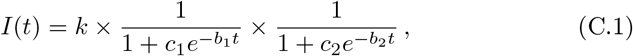

with

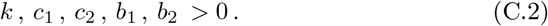

Before trying to fit *I* to some time series, one needs to resolve the degeneracy related to the symmetry (*b*_1_*,c*_1_) ↔ (*b*_2_*,c*_2_), This can be removed by requiring that

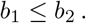

The case *b*_1_ = *b*_2_ would have required special attention, but in all the fits considered this case has never occurred, so we will from now on assume

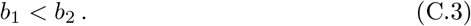

One can write *c*_1_ as 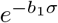 and *c*_2_ as 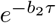, which brings (C.1) to the form

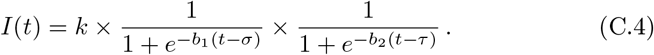

The initial growth rate of *I* is *α*:= *b*_1_ + *b*_2_, while the final extinction rate is

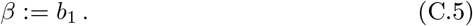

Renaming thus *b*_1_ to *β* we can write

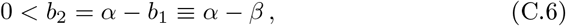

which leads to the following form of (C.4):

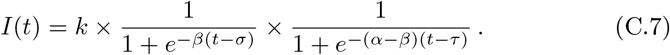

The first inequality in (C.6) shows that *α*>*β*: equivalently, a necessary condition for *I* to provide a good description of the epidemic the initial growth rate must be larger than the final extinction rate. We will see shortly that in fact we must have the inequality (C.8).

We emphasise that the above renaming of the exponents *b*_1_ and *b*_2_ requires that the exponent which has been renamed to *β* should be smaller than the exponent that has been renamed to *α* − *β*. Hence, if a fit of the data to the function (C.7) has been made, and if the parameters so determined are such that the parameter which was thought to be *α* − *β* is smaller than the parameter which was thought to be *β*, then the parameter which has been renamed to *α* − *β* should actually be *β*, and the parameter that has been renamed to *β* should be *α* − *β*.

This somewhat confusing discussion leads to a more stringent restriction for *I* to provide a good fit to the data: Since *β* ≤ *α* − *β* we must have

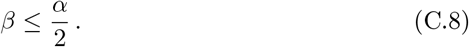

This shows that no good fit to the data can be found if (C.8) fails.

The above is closely related to the question of uniqueness of the parameters in (C.7). Here the question is, for what sets of parameters (*α, β, σ, τ*) and (*α′,β′,σ′,τ′*) it holds for all *t*

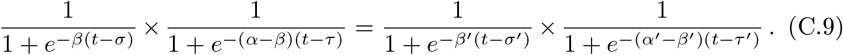

Equivalently,

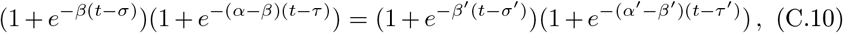

which is the same as

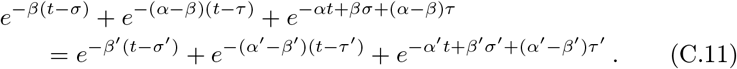

By comparing the fastest growing terms on both sides for *t* → −∞ one finds

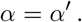

From the subleading exponential terms one obtains

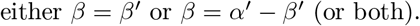

The first case leads to the obvious solution (*α, β, σ, τ*)=(*α′,β′,σ′,τ′*), while the second leads to

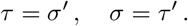

We have thus shown that the sets of parameters

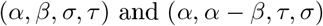

give the same function *I*, and that this is the only nontrivial possibility

In order to remove this degeneracy, within our fitting procedure we enforce the inequality

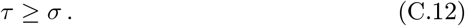

If a fit is found saturating the equality we repeat the procedure with the reverse inequality in place.

## Data Availability

All data available on the John Hopkins University server, https://coronavirus.jhu.edu

## Footnotes

1 This should be compared with the logistic function, where both the initial growth rate and the extinction rate are the same; the Gauss-error function, where both the growth and extinction rates vanish; and the Richards curve [2], where the initial and extinction rates are arbitrary but the growth amplitude is correlated with the extinction amplitude.

2 For example, if both *e*^(^*^α−β^*^)^*^τ^* ≥ 9 and *e^βσ^* ≥ 9, then the relative error in (1.2) less than 10%: 1*/x* − 1*/*(1 + *x*)=1*/*(*x*(1 + *x*)) ≤ 0.1 × 1*/x* if *x* ≥ 9. The accuracy of the approximation grows with growing *x*.

## Notes

### Competing Interest Statement

The authors have declared no competing interest.

### Clinical Trial

Research based on publicly available data.

### Funding Statement

No external funding

### Summary of Updates

Formatting of the supplementary material corrected

## References

[1] P. T. Chruściel, S. J. Szybka, *medRxiv:2020.05.22.20098350*.

[2] F. Richards, Jour. Exp. Botany 10, 290 (1959).

[3] L. Ferretti, et al., Science 368 (2020).

[4] X. He, et al., Nature Medicine 26, 672 (2020).

